# The Role of Fermented Pickles in Shaping Gut Microbiota and Immune Response in Women: A Community-Based Trial in Pakistan

**DOI:** 10.1101/2025.01.10.25320071

**Authors:** Sumbal Hafeez, Aqsa Khalid, Sheraz Ahmed, Fayaz Umrani, Abdul Khaliq Qureshi, Kumail Ahmed, Fariha Shaheen, Aneeta Hotwani, Furqan Kabir, Sean R. Moore, Syed Asad Ali, Junaid Iqbal, Najeeha Talat Iqbal

**Affiliations:** Aga Khan University, Karachi, Pakistan; Department of Cellular and Molecular Pathology, University of Wisconsin-Madison, Madison, WI, 53705, USA; Division of Gastroenterology, Hepatology, and Nutrition, Department of Pediatrics, Cincinnati Children’s Hospital Medical Center, Cincinnati, Ohio, USA

**Author notes:** Corresponding Author: **Najeeha Talat Iqbal**, Associate Professor, Address: Department of Pediatrics and Child Health, Department of Biological Biomedical Sciences, Aga Khan University, Stadium Road, Karachi 74800, Pakistan. Co-corresponding Author: **Junaid Iqbal**, Assistant Professor, Address: Department of Pediatrics and Child Health, Aga Khan University, Stadium Road, Karachi 74800, Pakistan.

## Abstract

A gut microbiome-targeted diet can potentially mitigate chronic diseases like malnutrition. In a prospective 12-week intervention trial, we evaluated the effects of six different plant-based fermented pickles (∼50g/day) on clinical, inflammatory, and gut-microbiome parameters in women (n=230) in a rural setting with a high prevalence of undernutrition. Blood was collected at two, whereas stool was collected at three timepoints. Among fecal biomarkers, myeloperoxidase (MPO), Lipocalin-2 (LCN2), and 16S rRNA sequencing were measured at baseline, 8^th,^ and 12^th^ weeks. Overall compliance rate was >70%. WBC and neutrophils significantly decreased among radish (p=0.002, p=0.01) and carrot (p=0.005, p=0.006) groups compared to controls. In lemon-chili groups, platelets significantly decreased (p<0.001) while MCV increased (p=0.02). In onion and lemon-chili groups, the alpha (р=0.001 and p=0.0005, respectively) and beta diversities (p=9e-04 and p=0.0223, respectively) were significantly increased. Post-intervention linear discriminant analysis (LDA) identified 25 bacterial taxa markers at 8^th^ and 12^th^ week, that included *Eggerthellaceae* and *Oscillospiraceae, Erysipelatoclostridiaceae* and *Subdoligranumlum,* predominantly in lemon-chili group. Correlation analysis revealed six taxa negatively associated with inflammatory markers such as CRP, LCN2, and platelets. Our study provides preliminary information about consumption of culturally acceptable fermented pickles exerting beneficial changes in hematological and gut microbiome profiles of women, post-intervention.

## Introduction

Malnutrition is becoming a severe global issue, particularly in low-middle-income countries (LMICs), affecting more than 2 billion people (1). The effect of fermented food on gut microbial changes is not fully elucidated, especially in areas of high malnutrition burden. In Pakistan women of reproductive age (WRA) mainly suffer from malnutrition due to inaccessibility to food, contaminated drinking water and limited choices of food. This often leads to disruption in gut homeostasis, dysbiosis of gut microbial species and increase in gut or systemic inflammation (2, 3). Fermented foods trials are limited to western world encompassing over the counter diary fermented foods (4), or non-traditional fermented food which do not reflect the diet of a rural malnourished women (5). Most studies highlight the beneficial effect of fermented food on gut microbial species, increasing diversity in microbial taxa post-treatment (4–7).

Microbiome-targeted diets profoundly influence the gut microbiome and immune biomarkers (8) such as mediterranean, vegan, or ketogenic diets that have been linked to gut microbiome modifications by activating innate and adaptive immune system (9–12). Microbial dysbiosis increases pathogenic bacteria in the gut and leads to immune system activation and inflammation (13, 14) whereas intestinal inflammation also releases myeloperoxidase and lipocalin, enzymes expressed by neutrophils for antimicrobial activity (15). Since, long- and short-term dietary interventions alter microbial phenotypes and the immune system, thus promising diets like fermented foods must be further explored in different populations to determine enhanced gut microbiome diversity and reduced inflammation, post-intervention (4, 16).

Plant-based fermented foods offer best solution of low-cost locally fermented products as a rich source of prebiotics such as complex carbohydrates, and probiotics like *Lactobacillus brevis*, *Pediococcus pentosaceus, Lactobacillus plantarum*, and *Lactobacillus fermentum*, which are favorable for gut and metabolic health (16–18). Fermentation converts complex carbohydrates and proteins into short-chain fatty acids (SCFA) like acetate, propionate, butyrate, and bioactive peptides respectively, improving nutritional value and bioavailability (19). Currently, plant-based fermented pickles like Kimchi, Jangajji, Sauerkraut, Torshi, Suancai, and traditional Chinese pickles, are mostly reported for their potential health benefits (20). South Asian countries have a long historical culture of consuming traditionally fermented and non-fermented pickles locally commonly known as “Achar” (21). Owing to the benefits of fermented pickle consumption, documented evidence in a clinical trial is much needed. The scarcity of human clinical trials, particularly, in malnourished settings of LMICs including Pakistan, necessitates investigation of the role of fermented foods in improving gut health and inflammatory biomarkers, particularly in women. In this clinical trial effect of daily consumption of 50 g fermented pickle was investigated on the gut health, clinical parameters (complete blood count (CBC) and C-reactive protein (CRP)), inflammatory biomarkers (myeloperoxidase (MPO), Lipocalin-2 (LCN2)), and 16S stool microbiome of women of reproductive in a resource-limited settings of Matiari, Sindh (22).

## Results

### Participants achieved up to 70% compliance with assigned fermented pickles during the intervention period

To examine the effect of pickle consumption on the gut microbiome and clinical parameters, 223 healthy women of reproductive age (WRA), ranging from 18 to 48 years, were recruited for an 8-week dietary trial (12-week complete protocol). Of 407 participants screened for the eligibility criteria, 223 were recruited and assigned to any of the 6 interventions and 1 control group who did not assign any treatment. Each arm had 30 participants. After 13 loss-to-follow-ups, the final enrollment was 29-31 per group (**Figure 1a**). The mean age of the women recruited was 30.2 ± 7.9 years (mean ± SD), with a mean BMI of 21.0 ± 4.1 kg/m² (mean ± SD). There was no difference in the baseline parameters such as Age, BMI and weight between groups. Predominantly, all participants were below 25 years of age and most of them were married (88%), and 8% were pregnant. The mean socioeconomic status among the groups was significantly different (p=0.02), highest in the mango-oil group (0.31± 0.7) and lowest in the carrot group (−0.3± 1.1). Among the baseline clinical parameters (BMI, weight, respiratory rate, temperature, blood pressure, heart rate), heart rate (p=0.03) and blood pressure (p=0.003) were significantly different. (**Table 1**).

**Figure 1:**
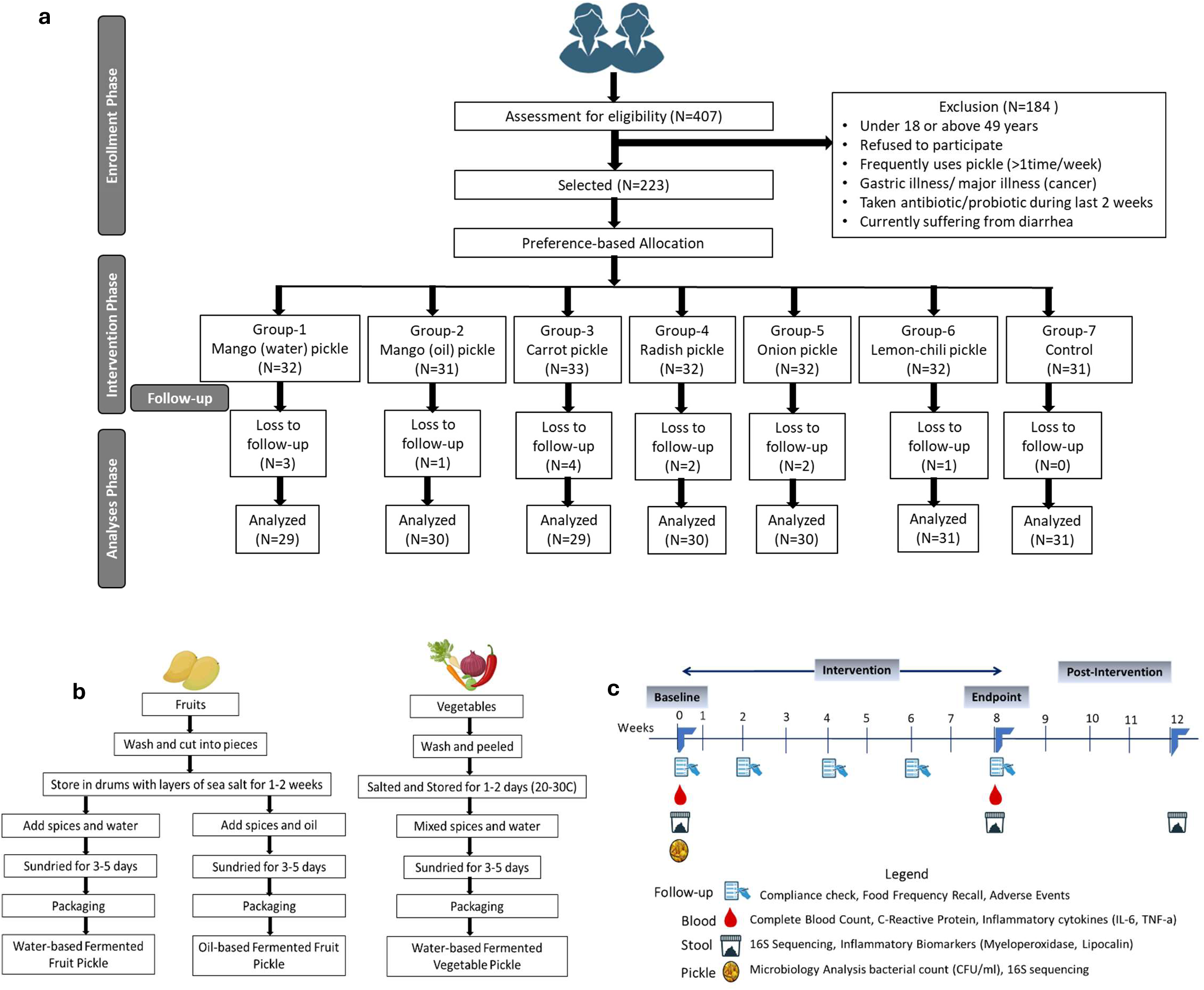
Overview of the fermented pickle intervention study. **(a)** Consort flow diagram for participant assessment, enrollment, allocation, follow-up, and analysis in each group. **(b)** Recipe followed for fermented pickle preparation. **(c)** The 12-week study timeline overview, sample collection, and experimental analysis.

**Table 1:** Demographic and Baseline characteristics of the participants enrolled in the study.

|  | Mango(water) | Mango (oil) | Carrot | Radish | Onion | Lemon | Control | *P* |
| --- | --- | --- | --- | --- | --- | --- | --- | --- |
| n | 32 | 31 | 33 | 32 | 32 | 32 | 31 |  |
| Age of participant | 28.4 ± 6.7 | 30.3 ± 8.0 | 32.2 ± 8.7 | 27.5 ± 7.1 | 26.7 ± 5.9 | 31.8 ± 7.5 | 34.3 ± 9.0 | 0.001* |
| Marital status |  |  |  |  |  |  |  |  |
| Married | 31 | 25 | 31 | 26 | 27 | 30 | 27 | 0.23 |
| Pregnancy status |  |  |  |  |  |  |  |  |
| Yes | 4 | 2 | 4 | 0 | 4 | 3 | 0 | 0.25 |
| SES (Wealth scores) | -0.2 ± 0.9 | 0.3 ± 0.8 | -0.3 ± 1.1 | -0.3 ± 1.0 | 0.1 ± 1.1 | 0.3 ± 1.1 | 0.1 ± 0.9 | 0.020* |
| Systolic pressure | 110.7 ± 10.1 | 109.3 ± 11.43 | 112.2 ± 11.6 | 116.7 ± 9.0 | 107.1 ± 7.9 | 109.91 ± 10.04 | 109.3 ± 12.4 | 0.015* |
| Diastolic pressure | 72.9 ± 7.4 | 74.7 ± 6.64 | 79.8 ± 8.9 | 77.7 ± 7.7 | 73.2 ± 8.5 | 74.91 ± 6.20 | 77.3 ± 9.6 | 0.003* |
| Weight (kg) | 45.0 ± 6.9 | 48.7 ± 10.88 | 49.4 ± 7.6 | 45.7 ± 9.7 | 49.7 ± 10.0 | 51.77 ± 13.17 | 46.2 ± 9.6 | 0.06 |
| BMI (kg/m<sup>2</sup>) | 20.3 ± 3.0 | 21.7 ± 4.5 | 21.1 ± 3.2 | 20.1 ± 4.1 | 21.7 ± 4.1 | 22.03 ± 5.21 | 20.2 ± 4.1 | 0.24 |
| BMI categories (kg/m<sup>2</sup>) |  |  |  |  |  |  |  |  |
| Underweight (<18.5) | 13 (40.6%) | 9 (29.0%) | 9 (27.3%) | 14 (43.7%) | 9 (28.1%) | 7 (21.9%) | 13 (41.9%) | 0.59 |
| Normal (18.5–24.9) | 12 (37.5%) | 12 (38.7%) | 15 (45.4%) | 14 (43.7%) | 12 (37.5%) | 16 (5%) | 14 (45.2%) |  |
| Overweight (25–29.9) | 3 (9.4%) | 3 (9.7%) | 6 (18.2%) | 1 (3.1%) | 6 (18.7%) | 4 (12.5%) | 3 (9.7%) |  |
| Obese (≥30) | 4 (12.5%) | 7 (22.6%) | 3 (9.1%) | 3 (9.4%) | 5 (15.6%) | 5 (15.6%) | 1 (3.2%) |  |

Data presented as mean ± SD and n (%). \**p* < 0.05. SES = Socioeconomic Status.

During our study, the participants successfully completed an 8-week intervention of assigned pickles, which was confirmed by measuring the weight of the pickle jar before handing it over to the participant and on return after 3 days. The compliance was categorized into 100%, 99-90%, 89-80%, and 79-70%. Among all groups, the carrot group participants showed the highest 100% compliance (93%) followed by radish (80%), mango-water (80%), mango-oil (71%), onion (47%), and lemon-chili (26%) groups. However, even the least compliant participants were above 70% compliant with the assigned intervention (**Figure S1**). A complete food recall log was maintained every 2 weeks follow-up and classified as cereals, fruits, vegetables, dairy products, meat, beverages, and pickles. The diet other than pickles was balanced among the groups with exception of meat which was less among the onion and lemon-chili participants (7% and 13% respectively) (**Table S1a-c**). Among side effects noted with Achar intervention were mostly abdominal discomfort and bloating etc. The majority of those were recorded for the radish group (28%) followed by lemon-chili (19%), and carrot (15%) (**Table S2**).

### Fermented pickle consumption modulates clinical parameters among intervention groups participants

The analyses of blood samples at week 0 and week 8 among the different intervention groups reported a significant modulation in a few clinical parameters. The radish group participants reported a significant decrease in mean white blood cells (Mean pre= 7.6×10E9/L; post=6.7×10E9/L, p=0.002), platelets (Mean pre= 316×10E9/L; post=283×10E9/L, p=0.01), and neutrophil (Mean pre= 57.6%; post=53.9%, p=0.01) counts post-intervention. Similarly, the carrot group participants also reported decreased platelet (Mean pre= 335×10E9/L; post=297×10E9/L, p=0.005) and neutrophil (Mean pre= 60.2%; post=56.2%, p=0.006) counts post-intervention. However, there was a significant decrease in platelets (Mean pre= 298×10E9/L; post=232×10E9/L, p=<0.001) and increase in mean corpuscular hemoglobin (MCH) (Mean pre= 26pg; post=27pg, p=0.02) and mean corpuscular hemoglobin concentration (MCHC) (Mean pre= 30.8pg; post=31.6pg, p=<0.001) among the participants of lemon-chili group, post-intervention. Among inflammatory biomarkers, LCN-2 increased (Mean pre= 46.9ng/ml; post=75.5ng/ml, p=0.01) only upon mango-water pickle intake. (**Figure 2 and Table S3a**).

**Figure 2:**
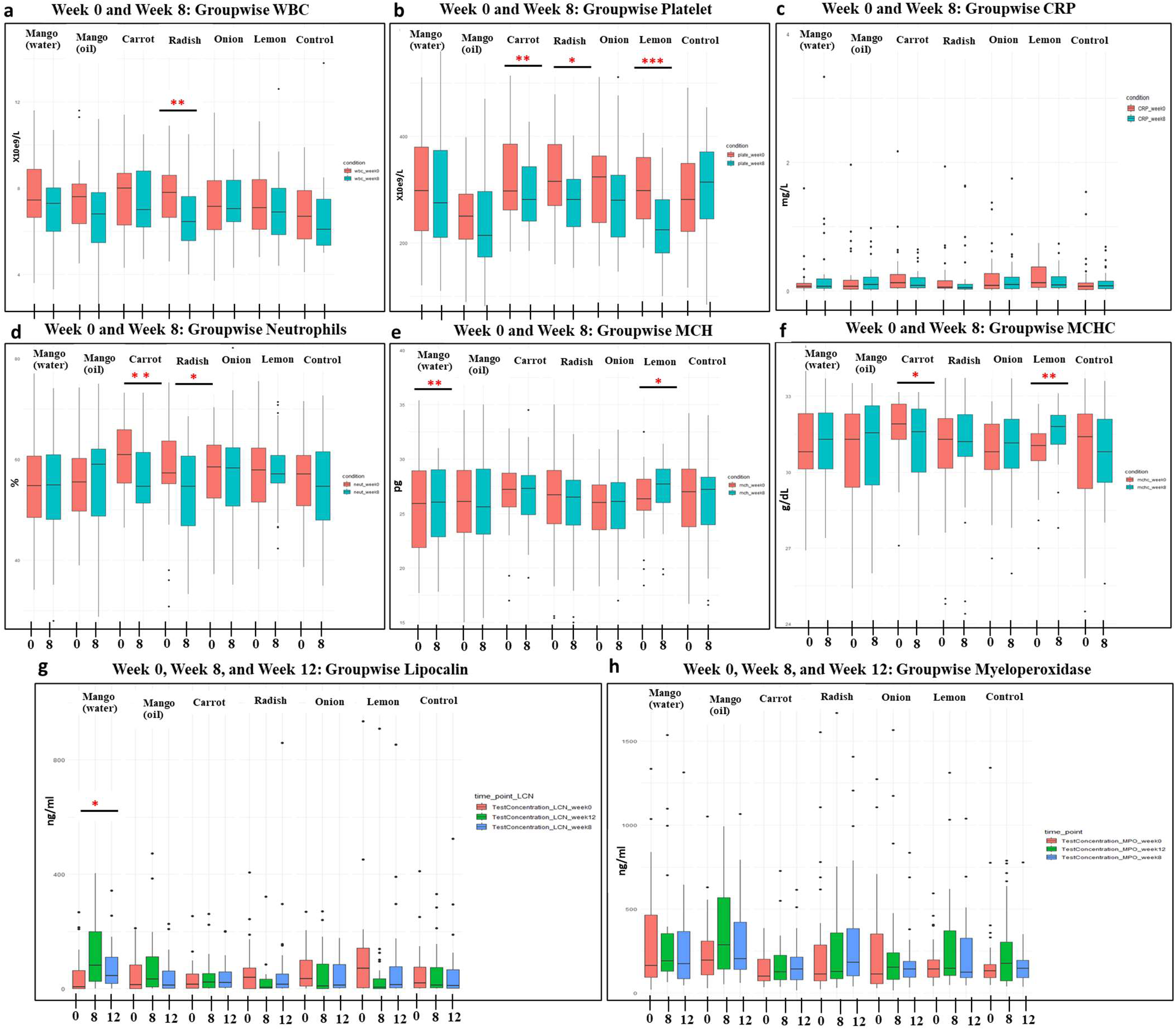
**(a-f)** Boxplot for week 0 and week 8 clinical biomarkers. **(a)** Radish participants WBC (p=0.002), **(b)** Carrot participants Platelets (p=0.005), Lemon-chili participants Platelets (p<0.001), **(d)** Carrot participants Neutrophils (p=0.006), Radish participants Neutrophils (p=0.01), **(e)** Lemon-chili participants Mean Corpuscular Hemoglobin (p=0.02), **(f)** Lemon-chili participants Mean Corpuscular Hemoglobin Concentration (p<0.001). (g-h) Boxplot for Week 0, Week 8, and Week 12 stool inflammatory biomarkers. (g) Mango-water participants LCN2 (p<0.01). * Represents a significant P-value by paired T-test or ANOVA.

In the multivariate model adjusted for fermented foods consumption other than pickles and baseline parameter taken as reference, there was a decreasing trend in WBC among all intervention groups, but significant reduction was noted among the radish group participants (ß= −0.92, CI= −1.7, −0.13, р<0.05). Similarly, the platelet counts were decreased among all the intervention groups but the reduction among the lemon-chili group was significant (ß= −65.9, CI=-100.3, −31.6, р<0.05) (**Table S3B**).

### The presence of culturable bacterial strains in the consumed pickles

Microbiological analyses of MRS and BHI media showed the presence of culturable bacteria in the pickles (**Figure S2**). Bacterial Sequencing of the selected gram-positive strains identified lactic acid bacteria (LAB) including *Lactiplantibacillus Pentosus, Lactiplantibacillus plantarum, Pediococcus pentosaceus, Pediococcus acidilactici, Levilactobacillus Brevis,* and *Weissella cibaria* and *Staphylococcus hominins* in the pickles (**Table S4**).

### Fermented pickle consumption remodeled the 16S stool Microbiome

Overall, 631 stool samples were collected from the enrolled participants. The average read count was 45,102 paired-end reads. 585 bacterial taxa were detected, however with a relative abundance >2.5% only 10 taxa in Intervention group and 13 in control group were identified (**Figure S3**). The core taxa of the intervention and control groups were consistent, with differences in the relative abundance of species including *Bifidobacterium, Prevotella, Collinsella, and Catenibacterium* (**Figure S4**). Alpha and beta diversities were similar across the group, post-intervention (**Figure S5-S8**). Since gut microbiome greatly varies at individual level, the parallel-design study comparison between intervention and control group is avoided to reduce the effect of individual variability (23). We therefore made comparisons within each group by analyzing the pre- and post-intervention gut microbiomes within intervention groups.

#### Pre- and Post-Intervention Alpha Diversity

The calculated “Observed” and “Shannon” α-diversity indices showed significant variations across the intervention groups. The Observed α-diversity among the mango-water (р<0.001) and carrot groups (p=0.02) changed significantly by decreasing at week 8 and increasing at week 12. Whereas the observed α-diversity of the onion group significantly increased at week 8 and week 12 (р=0.001). Similarly, both observed (р=0.0005) and Shannon (p=0.04) diversity indices increased significantly among the lemon-chili group at both week 8 (end-intervention) and week 12 (post-intervention) time points (**Figure 3a-f** and Figure S9).

**Figure 3:**
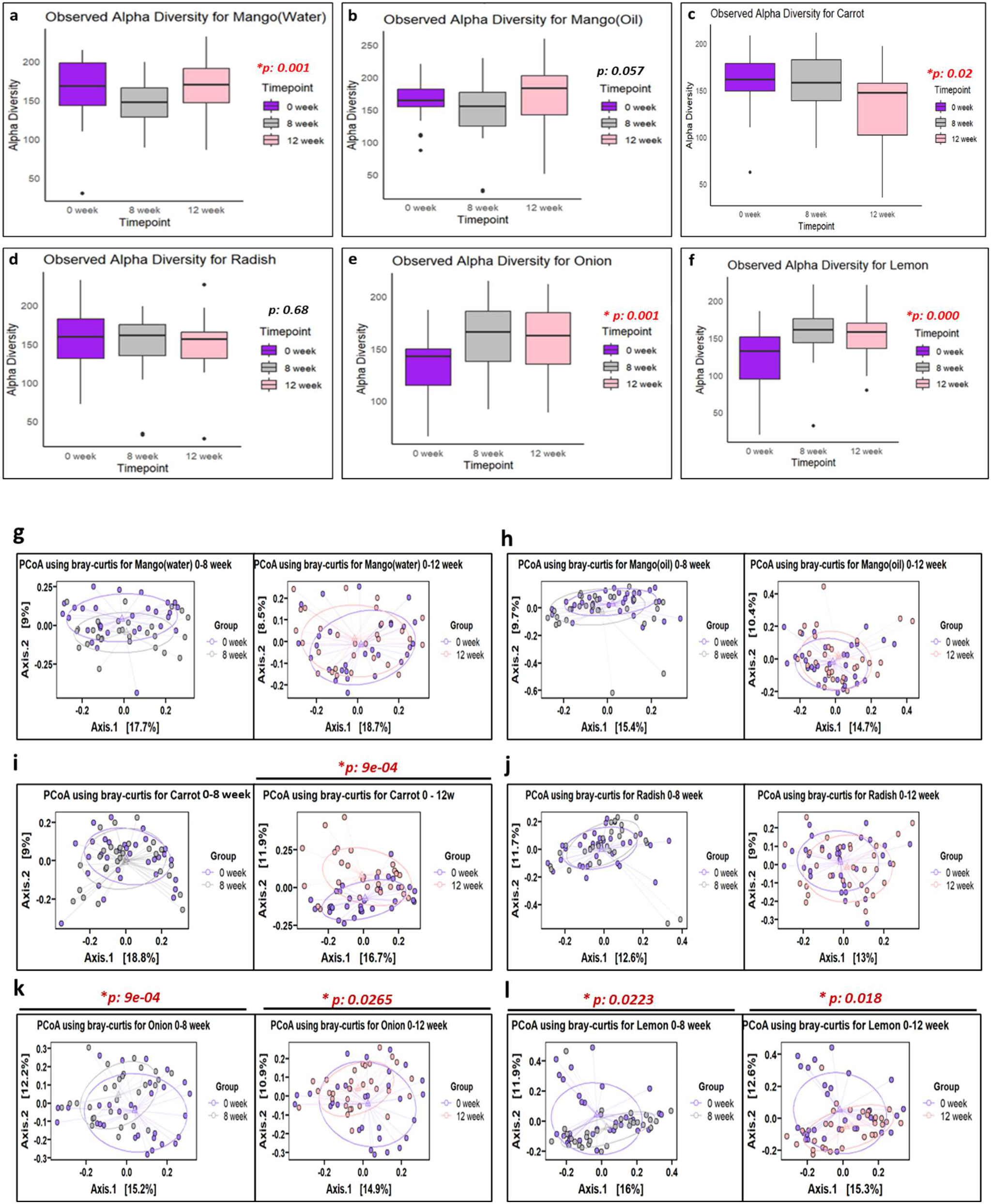
**(a-f)** Alpha-diversity of participants in intervention groups. **(g-l)** Beta diversity of the participants at 0-8 weeks and 0-12 weeks in intervention groups.

#### Pre- and Post-Intervention Beta Diversity

Beta diversity as calculated pre- and post-intervention did not change significantly among the participants of the mango-water, mango-oil, and Radish groups both at week 8 and week 12 when compared to pre-intervention. Among six interventions, β-diversity which is a measure of within sample variation showed significant change between week 0 to week 8 of the onion (p=9e-04) and lemon-chili (p=0.02) groups. Similarly, there was a significant variation between week 0 to week 12 among onion (p=0.0265) and lemon-chili (p=0.018). The β-diversity of the participants in carrot pickle did not change at week 8 but significantly changed at week 12 (p=9e-04) which shows that gut adaptation among the carrot pickle participants took a longer period than the onion and lemon-chili participants (Figure 3g-l).

#### Pre- and Post-Intervention Composition and Relative Abundance of Bacteria

Based on the taxa rank counts, a relative abundance above 2.5% was plotted to analyze the change in bacterial communities over time in different groups. The Phylum-level abundance plot showed the presence of *Firmicutes*, *Bacteriodota*, and *Actinobacteriota.* Post-intervention, most of the intervention groups such as carrot, lemon-chili, mango-oil, and mango-water groups showed an increase in *Firmicutes* except for onion and radish groups where *Actinobacteriota* increased compared to the baseline bacterial abundance (Figure 4a). At the family level, the relative abundance above 2.5% belonged to *Ruminococcacea, Lachinospiraceae, Prevotellaceae, Eggerthellaceae, Coriobacteriaceae, Bifidobacteriaceae*, *Atopobiaceae*, and *Erysipelatoclostridiaceae.* The relative abundance of species from *Lachnispiraceaea, Bifidobacteriaceae,* and *Ruminococcaceae* families increased after fermented pickles intervention (Figure 4b). The Genus-level relative abundance plot above 2.5% included *Prevotella, Collinsella, Fecalibacterium, Catenibacterium, Blautia, Bifidobacteria,* and *Agathobacter* (**Figure S10**)

**Figure 4:**
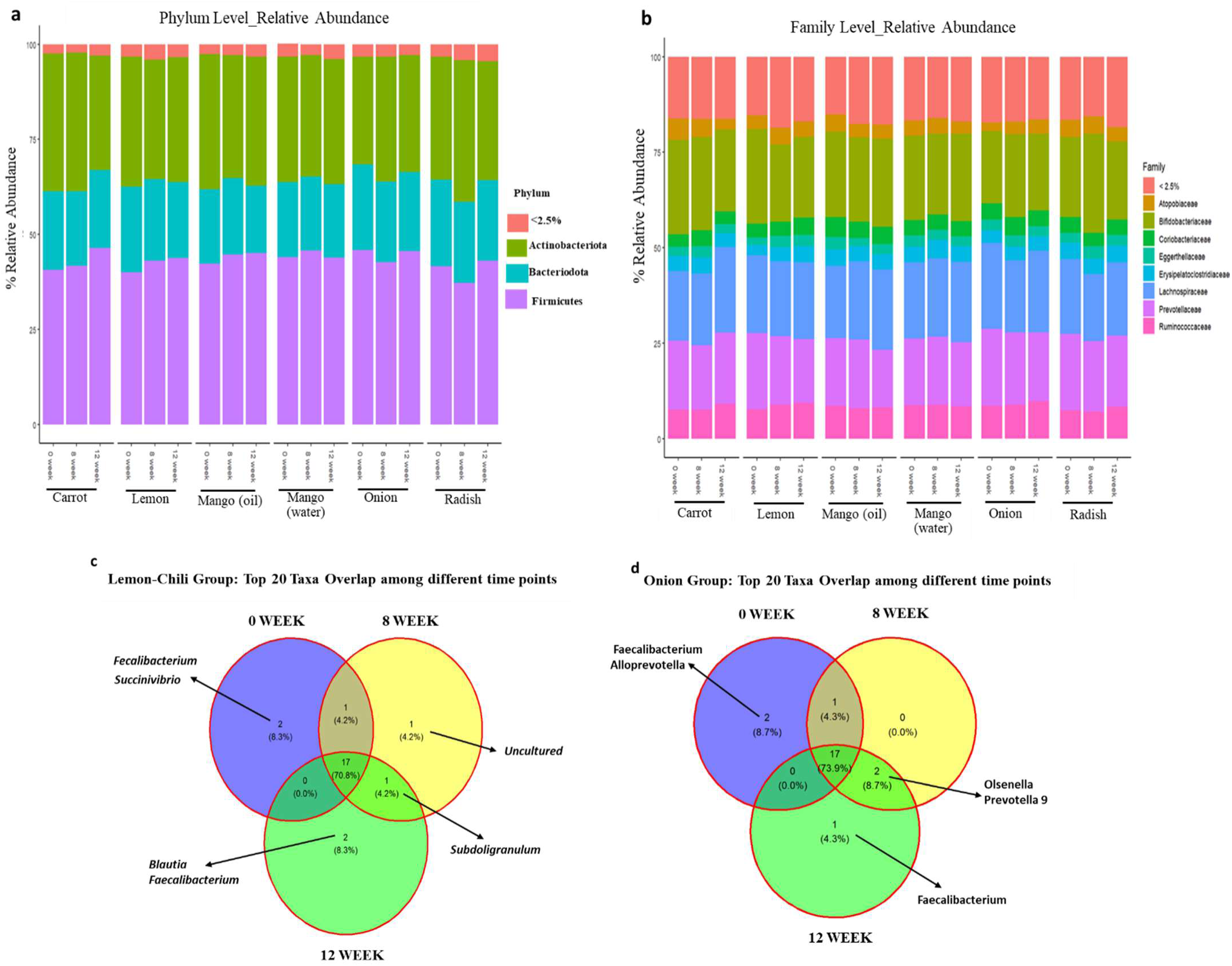
**(a)** Relative abundance of phylum among different intervention groups. **(b)** Relative abundance of family among different intervention groups. **(c)** Venn-diagram for shared top 20 taxa at three time points in the Lemon-Chili pickle intervention group participants. **(d)** Venn-diagram for shared top 20 taxa at three time points in the Onion pickle intervention group participants.

#### Pre- and Post-intervention Top-Taxa Remodeling

To identify unique bacterial taxa and persistence of bacterial taxa post-treatment, we plotted a Venn diagram to examine remodeling of the top 20 bacterial ASVs over time. The Venn diagram showed the shared and unique taxa at 3 time points. As the alpha and beta diversities significantly changed in the lemon-chili and onion group participants, we therefore focused on these two groups. The Venn diagram showed that approximately 70-75% of taxa were shared at 3 time points whereas only 25-30% differed post-intervention in both groups. The lemon-chili group had *Fecalibacterium* that appeared on week 12. However, the *Subdoligranulum* appeared at week 8 and persisted at week 12, even after the intervention was discontinued (Figure 4c). Similarly, in the onion group participants, *Olsenella* and *Prevotella* persisted both at week 8 and week 12 whereas there were no unique taxa at week 8. *Fecalibacterium* appeared in week 12 when the intervention was discontinued (Figure 4d).

#### Pre- and Post-intervention Linear Discriminant Analysis

Linear Discriminant Analysis (LDA) was plotted to examine the post-intervention significant bacterial markers. The results of significant bacterial markers were ranked by LDA scores at 3 timepoints. In the lemon-chili group, 25 bacterial taxa were significantly distinct including *Eggerthellaceae* and *Oscillospiraceae* with the highest LDA score at week 8 whereas 10 bacteria including *Erysipelatoclostridiaceae* and *Subdoligranumlum* showed the highest LDA score at week 12 (Figure 5a). Similarly, among the participants of the onion group, only *Actinobacteria* was significant marker of week 8 whereas *Olsenella, Singuinis*, *and Intestinibacter* were highly discriminant at week 12 (Figure 5b). The cladograms were plotted to show the phylogenic distances of the distinct taxa in both groups. The diameter of circle represented the abundance of the taxon (Figure 5c-d). In the mango-oil group, *Agathobacter*, *Eubacterium*, and *Dorea* were distinct at week 8 and week 12, respectively. In contrast, Dorea was highly distinct in the mango-water group at week 12 (**Figure S11a-b**). In the carrot group, *Coriobacteriales* and *Colinsella* were distinct at week 8 and *Firmicutes, Clostridia, and Lachnospiraceae* at week 12 whereas in the radish group, *Firmicutes* and *Eggerthellaceae* were discriminant taxa at week 12 (**Figure S11c-d**).

**Figure 5:**
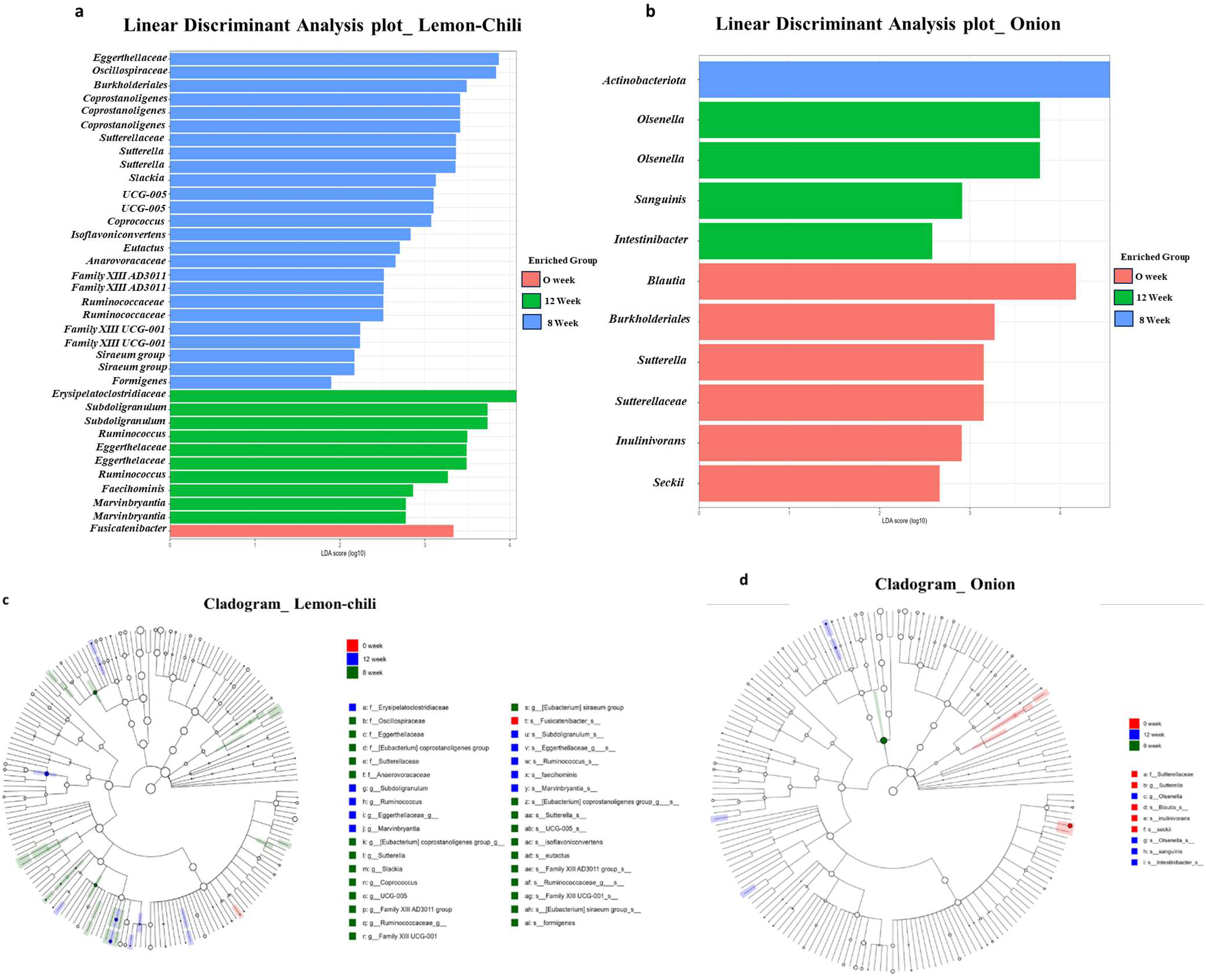
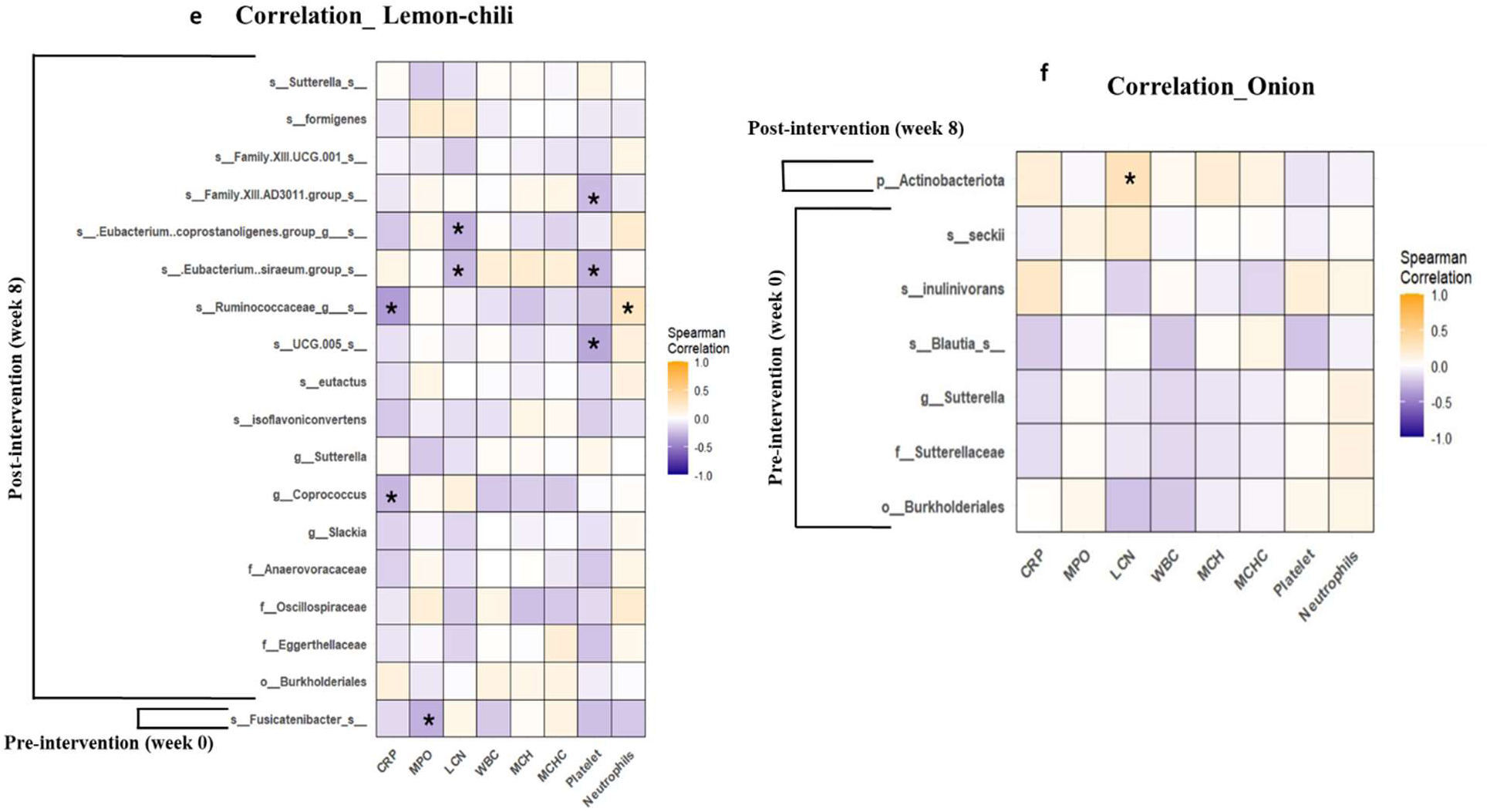
**(a)** LDA for the lemon-chili pickle intervention group participants at three time points. **(b)** LDA for the Onion pickle intervention group participants at three time points. **(c)** Cladogram of distinct taxa in the lemon-chili pickle intervention group participants. **(d)** Cladogram of distinct taxa in the onion pickle intervention group participants. **(e)** Correlation analysis of pre (week 0) and post (week 8) distinct taxa with clinical biomarkers in the lemon-chili pickle intervention group. **(f)** Correlation analysis of pre (week 0) and post (week 8) distinct taxa with clinical biomarkers in the onion pickle intervention group participants. The taxa belonging to the same group with the same relative abundance were considered one. In heatmap, yellow squares indicate significant positive correlations (Rho > 0.5, *p* < 0.05) and blue squares indicate significant negative correlations (Rho < − 0.5, *p* < 0.05). \**p* < 0.05.

#### Pre- and Post-intervention Correlation Analysis between Gut-microbiome makers and Clinical variables

The spearman correlation analyses of distinct taxa identified at week 0 (pre-intervention) and week 8 (post-intervention) with clinical variables significant at univariate level was performed using R-studio. In lemon-chili group, *Fusicatenibacter* identified at week 0 was negatively correlated (rho=-0.3; p=0.01) to myeloperoxidase, while the taxa identified post-intervention did not correlate with myeloperoxidase. CRP was negatively correlated with *Ruminococcaceae* (rho=-0.3; p=0.03), and *Coprococcus* (rho=-0.4; p=0.00). Lipocalin was negatively correlated with *Eubacterium coprostanoligene* (rho=-0.3; p=0.02) and *Eubacterium siraeum* (rho=-0.3; p=0.03). *Oscillospiraceae (UCG005)* (rho=-0.3; p=0.01) and *Eubacterium siraeum* (rho=-0.3; p=0.02), and Anaerovoracaceae *(Family XIII AD3011)* (rho=-0.2; p=0.04). *Ruminococcaceae* (rho=+0.3; p=0.04) was positively correlated with neutrophils. In the onion group, only post-intervention *Actinobacteriota* (rho=+0.3; p=0.02) was positively correlated with lipocalin (**Figure 5e-f** and **Table S5a-b**).

## Discussion

As gut microbiota-targeted diets like fermented foods have the potential to improve the gut microbiome (8). Our study highlights the first community-based intervention trial of plant-based fermented food in Pakistan. This study was designed to overcome the barriers to the intervention of fermented food, such as availability, standardization of doses, affordability, accessibility, social acceptability as a food item, and variable consumption of diverse fermented food available over the counter. These factors limit the generalizability of the results of fermented food trials.

Most well-designed fermented food intervention trials are limited to the Western world and rare in highly malnourished settings, where such trials are the most required to overcome food insecurity and anticipated beneficial role in gut homeostasis. Plant based fermented foods have an added benefit of extended shelf-life with no specific requirement for refrigeration storage making it suitable for less-privileged population in rural settings. Such a diet is also suitable for lactose intolerants and vegans, and religious constraints related to animal-based food consumption. It is convenient to either prepare such traditional fermented foods at domestic level or scale-up as a local business to help women empowerment (24).

This current trial was conducted with the goal to assess the impact of different traditional plant-based fermented foods intake, prepared by a local women entrepreneur, on the health of women of reproductive age living in high malnourishment area. The main findings of this work highlight the significance of fermented food in ameliorating gut microbiota. The notable findings of our study are: 1) modulation of hematological parameters post-intervention; 2) identification of LABs in locally fermented achars, 3) change in alpha and beta diversity indices post-treatment, and persistence remodeling of gut microbiome at 8- and 12-weeks post intervention, and 4) identifying distinct bacterial markers and their correlation with clinical biomarkers, post-intervention.

In this 12-week trial, the critical examination of multiple time points parameters of women revealed that different fermented pickles consumption uniquely modulated the gut microbiome and modestly affected the clinical parameters. This study showed successful compliance with 50g/day fermented pickle intake by the study participants. The minimum compliance for all intervention groups was 70%, which showed that allocating intervention based on choice was an effective strategy for better compliance. Most participants consumed the assigned pickles daily, especially mango-water and mango-oil pickles. Some participants shared reservations about the taste of the carrot and lemon-chili pickles. Daily consumption and taste inclination make mango-water, the preferred pickle, followed by mango-oil, and onion. Abdominal discomfort, heartburn, and diarrhea were the frequently reported side effects, predominant among the radish group participants followed by the lemon-chili, and carrot groups. Similar side effects were reported by another clinical trial conducted on fermented vegetable intervention in Western women (5).

Upon assessing the impact of fermented pickle intervention on the gut microbiome diversities, this study reported that among 6 intervention groups, there was a profound effect of lemon-chili and onion pickle on modulating gut microbiome as evident by the changes in alpha and beta diversity indices. The results showed that upon regular onion and lemon-chili pickle intake for 8 weeks, the participant’s gut microbiome diversity significantly increased and sustained for at least 4 weeks, even after the discontinuation of intervention, as found at post-intervention follow ups. Fermented foods contain probiotics and related metabolites such as polyphenolic compounds and SCFA that potentially increase the diversity and functionality of gut microbiome (25). However, mango-water, mango-oil, radish, and carrot participants showed either non-significant or delayed changes in gut microbiome diversity. It could be due to a short intervention duration providing insufficient time for gut microbiome remodeling (26). Environmental factors could also hinder the gut microbiome diversity including inadequate diet due to socioeconomic factors, continuous exposure to pathogens, lack of access to clean water, or lack of sanitation and hygienic lifestyle (27, 28). The increase in gut diversity upon pickle consumption aligns with the previous prospective cohort study conducted in the Chinese population where pickle consumption significantly reduces the risk of diabetes by increasing the gut microbiome diversity (29). However, another study conducted on the Pakistani population associated pickle consumption with low alpha diversity, as this study found with mango-water and carrot pickle interventions (30). Hence, this study concluded that the type of vegetable pickle showed uneven effects on the gut microbiome diversity.

One of the aims of this study was to explore the impact of fermented pickles intake on clinical and inflammatory markers. The data showed that in the participants who consumed lemon-chili pickle, a significant reduction in platelets was observed at week 8. Platelets are known to interact with immune cells in inducing immune-mediated inflammatory diseases (IMIDs) (31). Other inflammatory biomarkers including WBCs, CRP, and myeloperoxidase, though insignificant, were also decreased among the lemon-chili group’s participants. Similarly, a significant reduction in WBCs was observed among the participants who consumed radish pickles. WBC elevation is also a well-known inflammation biomarker and a decrease in WBC count represents a recovery trajectory in active inflammation patients (32). However, among most of our other intervention groups including mango-water, mango-oil, carrot, and onion, insignificant modulations were reported in clinical and inflammatory parameters except for increase in lipocalin upon mango-water pickle intake. Lipocalin belongs to the family of proteins involved in inflammation and adaptive immune response, and increase in lipocalin could be due to enhanced bioavailability of bioactive compounds like carotenoids and polyphenol which are known to modulate inflammatory pathways (33, 34). The insignificant change in inflammatory parameters coincide with a 6-week clinical trial of fermented vegetable consumption in women where inflammatory biomarkers change did not reach statistical significance (5).

We also assessed gut microbiome composition among different group participants. Previously, it has been reported that diets with high-fat and sugar have low abundance of fiber-degrading taxa like *Prevotellaceae, Ruminococcaceae*, and *Lachnospiraceae* (35). We hypothesized an increased abundance in these taxa in fermented pickles intervention groups. The results of relative abundance data from the current study reported a notable shift in *Lachnispiraceaea, Bifidobacteriaceae,* and *Ruminococcaceae* families belonging to phylum *Firmicutes* and *Actinobacteria*, post-intervention. Most of these species belonging to phylum *Firmicutes* and *Actinobacteria* were also part of the core taxa of recruited women as identified at baseline. However, as suggested by a previous study, it is uncertain if the relative abundance of the already existing species increased or if new species were added with intervention (4). Though, an extensive study reported that fruit and vegetable-derived microbes can contribute to human gut diversity (36). The limited taxa isolated and identified from the pickles that were consumed by the participants included *L. Pentosus, L. plantarum, Pediococcus pentosaceus, Pediococcus acidilactici, L. Brevis,* and *Weissella cibaria* and *S. hominins* but these taxa differed from those found in the gut microbiome of the study participants. The similar taxa were reported by a review conducted on LAB isolated from Asian fermented foods (37).

Another goal of this study was to identify if the top-taxa remodeled post-intervention. Remodeling showed beneficial bacterial occurrence and persistence over time. The Venn diagram suggested that post-intervention, the majority of the top-taxa remained unchanged, increasing or adding a few taxa including *Subdoligranulum* and *Fecalibacterium* and *Prevotella* and *Olsenella* among lemon-chili and onion participants, respectively. Since these taxa persisted in both post-intervention time points, this shows the capability of the gut microbiome to adapt and sustain dietary changes (38). All these bacteria are SCFA producers, crucial for improved gut health and reduced inflammation (39, 40). The linear discriminant analysis further validates that different vegetable and fruit pickle consumption uniquely remodels the gut microbiome and presents distinct gut taxa among the participants. This could be attributed to different vegetables possessing dissimilar bioactive constituents (41) even if fermented under the same process and contain similar bacteria. As reported by Kiczorowski et al., the fermentation of different vegetables showed the synthesis of diverse essential enzymes and other active constituents including minerals, vitamins, antioxidants, phenols, and heavy metals (42).

Finally, this study also correlated the pre- and post-intervention distinct taxa identified by LDA analyses with significant clinical biomarkers. The analyses revealed that post-intervention taxa negatively correlated with lipocalin, CRP, and platelets suggesting the anti-inflammatory role of the intervention and predicts potential beneficial modulation in systemic inflammation through dietary changes (43). Conversely, positive association of bacterial taxa with neutrophils suggests enhanced immune response upon lemon-chili intake. This dual relationship of gut microbiome with immune system upon lemon-chili consumption highlights the complex interaction of gut microbiome, diet, and host health (44).

Rich distinct taxa and its correlation with clinical biomarkers in the lemon-chili intervention group favors the hypothesis that combining the beneficial components of different vegetables synergistically enhances gut diversity and functionality. This could be potentially due to combined effect of capsaicin, an active constitute of chili which increases the abundance of *Ruminocacocaeae* and *Lachnocpiraceae*, both butyrate-producing bacteria and pectin, an active dietary fiber of lemon, which leads to the production of SCFA via gut microbiome (45). Given the distinct response by fermented lemon-chili pickle, future exciting opportunities could be to analyze the mixed-vegetables pickle response in gut microbiome compared to individual pickle intake. Furthermore, additional studies could be conducted to strategically assess the mechanistic role of fermented pickle intake in alleviating different health conditions particularly malnutrition and gut-inflammatory diseases in murine models and large diverse population cohorts.

## Conclusion

In conclusion, the findings of the present study highlight that regular intake of different fermented pickles has an uneven yet beneficial impact on the gut microbiome and other important clinical parameters, potentially improving overall health outcomes in women of reproductive age. Among 6 intervention groups, there was a profound effect of lemon-chili and onion pickle on modulating gut microbiome as evident by the changes in alpha and beta diversity indices. However, future investigations on a large population are required to unravel the interaction of fermented pickles, particularly mixed pickles, on the gut microbiome and overall health outcomes. Additionally, murine models of malnutrition and gut inflammatory disease could be used to discover mechanistic insights into diet-microbiome interactions. Such findings will be useful in dealing with other noncommunicable chronic diseases.

## Materials and Methods

### Study design and site

A community-based multi-arm clinical trial was conducted in Matiari, a rural district of Sindh, Pakistan. 223 women of reproductive age (WRA) were recruited in the study designed to explore the impact of a fermented pickle diet on the gut microbiome and immune parameters. Approval from the Ethical Review Committee of Aga Khan University was obtained (ERC-2022-6595-23253: Grand Challenges Fermented Food - Achars (fermented pickles) in Pakistan). The trial was registered on <u>Clinicaltrials.gov</u>, identifier: NCT06748313. Informed consent was taken from all the recruited participants.

### Eligibility criteria

Participants under 18 or above 49 years of age, with any gastric/major illness history, frequently consumed pickles, or antibiotics/probiotics within 2 weeks of the interview date were excluded from the study.

### Fermented Pickle Preparation

Fermented pickles were prepared by a local vendor using a traditional recipe. The fresh vegetables and fruits (5 kg) separately, were cut and mixed with turmeric (50g), salt (100g), mashed mustard seeds (50g), chili powder (60g), spring garlic (250g), mustard oil (50ml), and water or oil (3L) – depending on water-based or oil-based achar. The prepared mixture was then put under shade at 20-30℃ for 2-14 days, sun-dried for 3-5 days, and packed for consumption. These recipes were aligned with those reported by Bhera et al. (46) with a few traditional changes (Figure 1b).

### Intervention

The participants were assigned to 6 different fermented pickles based on their preference, those who did not like pickles were assigned to the control group (30/per group, total N=210). The total number of participants enrolled was 223, to accommodate the loss-to-follow-ups. The intervention groups included the mango-water-based pickle group (N=32), mango-oil-based pickle group (N=31), carrot pickle group (N=33), radish Pickle group (N=32), onion Pickle group (N=32), and lemon-chili pickle group (N=32) and control group (N=31). All participants except the control group, consumed 50g/day of fermented pickles for 8 weeks in addition to their regular meals. Refilling of the pickles was ensured every third day. The food recall questionnaire, side effects, clinical parameters, and compliance follow-up data were collected every 2 weeks (Figure 1c).

### Microbiological analysis of food and bacterial sequencing

The pickle samples were cultured using De-mannose Ragosa Sharpe (MRS) Agar and Brain Heart Infusion (BHI) agar to calculate the number of live bacteria present in the pickle brine. The brine was serially diluted using 1X Phosphate Buffer Saline (PBS). Agar plates were incubated at 37℃ for 48-72 hours under aerobic and anaerobic conditions. The colony forming units (CFU) per ml were counted from a countable plate and colonies were sub-cultured to isolate pure colonies stored in 20% glycerol at −80℃ for later use.

DNA was extracted from the selected gram-positive colonies isolated using a lysis buffer and Qiagen DNA extraction kit (Qiamp-51106) according to the manufacturer’s instructions. Lysis buffer was prepared by adding lysozyme to a lysis solution containing 1M Tris-HCl (pH 8.0) (Sigma), 0.5M EDTA (pH 8.0) (Invitrogen), and Triton X-100 (Sigma-Aldrich). DNA was subjected to shallow sequencing for taxonomic identification of pure bacterial strains. Kit used for library preparation (DNA prep). The samples were multiplexed using an indexing kit and pooled to generate a single library loaded onto the MiSeq flow cell (v3 – 600 cycles kit). For every sample, reads with Q>=15 was used for taxonomic identification. The Bacterial and Viral Bioinformatics Resource Center (BV-BRC) platform was used to identify the taxonomy of bacteria using paired-end reads (https://www.bv-brc.org/).

### Compliance, Side Effects, and Food Recall Data

The food frequency recall questionnaire, side effects, and medication intake were logged by a trained research staff every 2 weeks. Food was classified as cereals, fruits and vegetables, dairy products, meat, beverages, and pickles. To avoid sharing weighed pickles, an additional jar was provided to the participants for other house members, and compliance was validated by measuring the weight of the jars returned. Compliance was calculated using the formula “Total achar consumed = weight of achar bottles provided – weight of achar bottles remaining”. Percentage compliance was calculated using following formula:

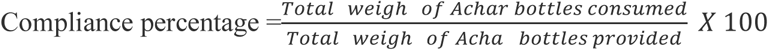

### Anthropometric measurements and clinical parameters

Anthropometric measurements, heart rate, temperature, respiratory pressure, and blood pressure were documented every 2-week follow-up during the intervention.

### Sample Collection

Stool samples were collected at week 0 (pre-intervention), week 8 (end-intervention), and week 12 (post-intervention) by the field team within 30 minutes of stool pass and snap-frozen using dry-ice for transport to the Matiari Research Lab and stored at −80 ℃.

Similarly, blood samples were collected at week 0 (pre-intervention) and week 8 (end-intervention) in EDTA tubes and transported to the MRL at 4 ℃. An aliquot of blood was used to measure complete blood count (CBC) and C-reactive protein (CRP) levels.

### Stool Inflammatory cytokines

Commercial ELISA kits were used to measure intestinal inflammation using a myeloperoxidase (MPO) kit (Immunodiagnostic AG, Stubenwald-Allee, Bensheim), and lipocalin (LCN) (GenWay Biotech, San Diego, CA, USA). All protocols were performed per the manufacturers’ instructions. The final dilution was measured by determining the appropriate concentration of a biomarker using a linear range of the standard curve. LCN at the dilution of 1: 30 and 1:50, and MPO at 1:50 and 1:100 was considered. All plates were read on 450 nm using a BioRad iMark (Hercules, CA) plate reader.

### Stool DNA extraction

DNA was extracted according to the manufacturer’s protocol, using a Qiagen DNeasy Power Soil Pro Kit (Qiagen, Germany) and quantified using Invitrogen Qubit 1X dsDNA HS assay kit (Thermo Scientific, USA) on a Qubit Fluorometer. The purity of DNA was assessed using a Nanodrop 2000 spectrophotometer (Thermo Scientific, USA) and stored at −80⁰C ULT freezer, until further use.

### 16S amplicon sequencing and statistical analyses

The DNA samples were shipped to the University of Minnesota Genomic Center (UMGC) for amplification of the V4 region of the 16S ribosomal RNA (rRNA) subunit gene (4). On average 45,102 reads per sample were generated. The quality of the raw fastq files was assessed using MultiQC and were further processed using the DADA2 pipeline to denoise (with the following parameters used in the filterAndTrim-step: maxN = 0, maxEE = c(2,2), truncQ = 2, rm.phix = TRUE, compress = TRUE), dereplicate reads, merge pair-end reads, and remove chimeras (47). Amplicon sequence variants (ASVs) were assigned using the naive Bayesian classifier via assignTaxonomy command and aligned against the SILVA rRNA database (https://benjjneb.github.io/dada2/training.html). A phylogenetic tree was constructed using the QIIME2“q2-phylogeny” plugin based on the ASV sequences. The data objects were combined into a Phyloseq object for downstream analysis. After pre-processing, the 585 ASVs were normalized by scaling the sample counts using a compositional transformation. This transformation ensured uniform representation of ASV counts across all samples, to facilitate effective comparison. The relative abundance of ASVs was calculated, and those with relative abundance >2.5% were considered as the most abundant within the bacterial community for each group.

Alpha diversity metrics (observed and Shannon) were measured using the plot richness function in Phyloseq with the significance calculated using the Kruskal-Wallis’s test. Beta diversity was calculated using the Bray–Curtis index with 9999 permutations (Vegan package) and visualized as principal component analysis (PCoA) ordination plots. Pairwise PERMANOVA test was conducted to identify significant differences among groups. Intergroup comparison among the top 20 highly abundant taxa were used to identify the common and unique taxa.

Linear discriminant analysis (LDA) effect size (LEfSe) analysis was used to determine the taxa contributing to the differences among the groups at 0, 8, and 12 weeks, with significance tested Kruskal-Walli’s sum-rank test (p <0.05), followed by LDA to estimate effect size at log (10) values. The LDA cut-off was set to 0.0 across all the groups. The cladograms were plotted using ggdendro package in R. The correlation of taxa markers from lefse (0 week and 8 week) with blood (WBC, MCH, MCHC, neutrophils) and inflammatory (CRP, MPO, LCN) parameters were calculated using spearman’s correlation. All the microbiome data analysis was performed using R v.4.10 software.

Variables other than microbiome data were analyzed using Stata/SE 17.0 for Windows. Continuous variables were summarized as means with standard deviations (mean ± SD), while categorical variables were reported as frequencies and percentages. A paired t-test for two timepoints and repeated measure Analysis of variance (ANOVA) for three timepoints were utilized to compare mean values across groups, with post-hoc tests applied for specific group comparisons. The chi-square test was used to assess differences in categorical variables between groups. Additionally, multivariate regression analysis was conducted to control potential confounding variables.

## Supporting information

List of Supplementary Figures

## Acknowledgments

We thank our research and field teams at Matiari and the study participants for complying with our study protocol.

## Author Contributions

The authors’ responsibilities were as follows: NTI,JI,SA,SAA: designed research; SH,SA,KA,AH,FK: conducted research; AK,SH: analyzed data; SH: wrote the paper; SAA,NTI: had primary responsibility for final content; FS: provided data management; SH,AK: provided data interpretation; NTI,SRM,SAA: provided substantial manuscript editing

## Authors Approval

All authors read and approved the final manuscript.

## Funding

This study was funded by the Bill & Melinda Gates Foundation (BMGF) through a grant: Global Grand Challenges: Integrating Tradition and Technology for Fermented Foods for Maternal Nutrition (INV-033567) https://gcgh.grandchallenges.org/grant/achars-pickles-reduce-inflammation-and-improve-microbiome-rural-pakistani-women. SH received research training support from the National Institute of Health’s Fogarty International Center (5D43TW007585-13). The funders had no role in the design, data collection, analysis of the study, or the decision to publish or prepare this manuscript.

## Data availability

All de-identified metadata and microbiome data related to this study will be available after publication. Please contact corresponding author, NTI at for data request.

## Conflict of interest

All authors have no conflict of interest to declare.

## Notes

### Competing Interest Statement

The authors have declared no competing interest.

### Clinical Trial

NCT06748313

### Clinical Protocols

https://clinicaltrials.gov/search?cond=NCT06748313

https://gcgh.grandchallenges.org/grant/achars-pickles-reduce-inflammation-and-improve-microbiome-rural-pakistani-women

### Author Declarations

Approval from the Ethical Review Committee of Aga Khan University was obtained (ERC-2022-6595-23253: Grand Challenges Fermented Food - Achars (fermented pickles) in Pakistan)

