## Supplementary material for "The Role of Fermented Pickles in Shaping Gut Microbiota and Immune Response in Women: A Community-Based Trial in Pakistan": List of Supplementary Figures

Supplementary Figures

Figure S1: a) Overall study Percentage Compliance of the intervention. b) Group-wise Participants Percentage Compliance of the intervention.

Figure S2: Culturable bacteria in fermented pickles (log CFU/ml) isolated on Brain Heart Infusion (BHI) agar under aerobic and De-Mannose Ragosa Sharpe (MRS) agar under aerobic and anaerobic conditions.


Figure S3: Overall highly abundant taxa in recruited women participants a) Controls b) Intervention.


Figure S4: Core taxa of recruited women participants: a) Control b) Intervention.

Figure S5: Pre-intervention (Week 0) alpha (Shannon) and beta-diversities between control and intervention groups.

Figure S6: End-of-intervention (Week 8) alpha diversity between cases and intervention groups.

Figure S7: Post-intervention (Week 12) alpha diversity between cases and intervention groups.

Figure S8: Beta-diversity at 3-time points between cases and intervention groups.

Figure S9: (a-f) Shannon alpha-diversity of the participants in intervention groups.

Figure S10: Relative abundance of genus among different intervention groups

Figure S11: Linear Discriminant Analysis intervention group participants at three time points (a) Mango (Water) pickle (b) Mango (Oil) pickle (c) Carrot pickle (d) Radish pickle.
